# Unraveling COVID-19-related hospital costs: The impact of clinical and demographic conditions

**DOI:** 10.1101/2020.12.24.20248633

**Authors:** Anna Miethke-Morais, Alex Cassenote, Heloísa Piva, Eric Tokunaga, Vilson Cobello, Fábio Augusto Rodrigues Gonçalves, Renata Aparecida dos Santos Lobo, Evelinda Trindade, Luiz Augusto Carneiro D‘Albuquerque, Luciana Bertocco de Paiva Haddad, on behalf of HCFMUSP Covid-19 Study Group

**Affiliations:** Clinical Director’s Office, Hospital das Clínicas, Faculty of Medicine, University of São Paulo, SP, Brazil; Department of Gastroenterology, Faculty of Medicine, University of São Paulo, São Paulo, SP, Brazil; Discipline of Labour Market and Physician’s Health, Santa Marcelina Faculty, São Paulo, SP, Brazil; Medical Demography Study Group, Department of Preventive Medicine, Faculty of Medicine, University of São Paulo, São Paulo, Sp, Brazil; Executive Director’s Office, Instituto Central, Hospital das Clínicas, Faculty of Medicine, University of São Paulo, SP, Brazil; Strategy and Operations Department, Hospital das Clínicas, University of São Paulo Medical School - FMUSP, São Paulo, Brazil; Information Technology Department, University of São Paulo School of Medicine FM-USP, São Paulo, Brazil; Affiliated Researcher, Health Technology Assessment Center of the Clinical Hospital (HC- FMUSP), São Paulo University Medical School (FMUSP), São Paulo, Brazil; Laboratory of Medical Investigation in Pathogenesis and Targeted Therapy in Onco-Immuno- Hematology (LIM-31), University of São Paulo Medical School - FMUSP, São Paulo, Brazil; Health Technology Assessment Center of the Clinics Hospital of the São Paulo University Medical School, NATS-HCFMUSP; Health Technology Assessment Center of the Executive Direction – Heart Institute, InCor- HCFMUSP; São Paulo State Health Technology Assessment Network, São Paulo State Health Secretariat, REPATS-SES/SP; Digestive Organs Transplant Division, Gastroenterology Department, University of São Paulo School of Medicine FM-USP, São Paulo, Brazil

## Abstract

**Introduction:** Although patients’ clinical conditions were previously shown to be associated with coronavirus disease 2019 (COVID-19) severity and outcomes, their impact on hospital costs is not known. The economic evaluation of COVID-19 admissions allows the assessment of hospital costs associated with the treatment of these patients, including the main cost components and costs driven by demographic and clinical conditions. The aim of this study was to determine the COVID-19 hospitalization-related costs and their association with clinical conditions.

**Methods:** Prospective observational cohort study of the hospitalization costs of patients with COVID-19 admitted between March 30 and June 30, 2020, who were followed until discharge, death, or external transfer, using micro-costing methodology. The study was carried out in the Central Institute of the Hospital das Clinicas, affiliated with the Faculty of Medicine of the University of Sao Paulo, Brazil, which is the largest hospital complex in Latin America and was designated to exclusively admit COVID-19 patients during the pandemic response.

**Results:** The average cost of the 3,254 admissions (51.7% of which involved intensive care unit (ICU) stays) was US$12,637.42. Overhead cost was the main cost component, followed by daily fixed costs and drugs. Sex, age and underlying hypertension (US$14,746.77), diabetes (US$15,002.12), obesity (US$18,941.55), cancer (US$10,315.06), chronic renal failure (US$15,377.84), and rheumatic (US$17,764.61), hematologic (US$15,908.25) and neurologic diseases (US$15,257.95) were significantly associated with higher costs. Age >69 years, RT-PCR-confirmed COVID-19, comorbidities, the use of mechanical ventilation, dialysis, or surgery, and poor outcomes remained significantly associated with higher costs after model adjustment.

**Conclusion:** Knowledge of COVID-19-associated hospital costs and their impact across different populations can aid in the development of a generalizable and comprehensive approach to hospital preparedness, decision-making and planning for future risk management. Determining the disease-associated costs is the first step in evaluating the cost-effectiveness of treatments and vaccination programs.

**SUMMARY BOX:** *Question:* What are the COVID-19 hospitalization-related costs?

*Findings:* In this prospective cohort that was carried out in a single reference quaternary center designated for the treatment of severe cases of COVID-19, more than three thousand patients were included, and their costs of hospitalization were found to be directly related to the age and comorbidities. The costs were more than 50% higher in older patients, 10-24% higher in patients with comorbidities, and 24-200% higher when additional therapeutic procedures were required.

*Meaning:* Determining the disease-associated costs is the first step in conducting future evaluations of the cost-effectiveness of treatments and vaccination programs, supporting their implementation with a comprehensive population-based approach.

## INTRODUCTION

The economic impact on health systems worldwide is a major concern related to the COVID-19 pandemic, and there is an emergent need for additional resources and financial investments. The available hospital capacity, including hospital facilities, equipment, supplies and health professionals, has had to be significantly increased. Economic evaluations are thus essential to ascertain the health care-related resources and costs required for patients with this new disease.[1]

Available data suggest that 5–20% of patients with COVID-19 require hospitalization, and between 14 and 20% of them require intensive care unit (ICU) admission.[2] Underlying medical conditions and patient characteristics have already been associated with disease severity and outcomes. Comorbidities also affect the resource and treatment requirements because they can result in longer hospitalization periods, mechanical ventilation or dialysis, among others.[2-4]

In Latin America, the Hospital das Clínicas of the University of São Paulo Medical School (HCFMUSP) is the largest public hospital complex, with 2,400 beds, and was located in the national epicenter of the pandemic. The Central Institute of HCFMUSP, with 900 beds, including 300 ICU beds, 206 of which were newly installed in response to this pandemic, was entirely dedicated to COVID-19 patients referred for admission, and has delivered tertiary hospital care for more than 4,000 severe COVID-19 cases.[5-7] The centralization of care in a single institute provided a unique opportunity for a precise and homogeneous economic evaluation of these admissions. The aim of this study was to describe the direct and fixed hospital costs related to the treatment of hospitalized patients with COVID-19, as well as the main cost components and related clinical factors.

## METHODS

### Study design, inclusion and exclusion criteria and ethical aspects

This was an economic prospective observational cohort study conducted at the HCFMUSP. All consecutive patients admitted from March 30 to June 30, 2020 with suspected, probable or confirmed COVID-19[8, 9] were included and followed until discharge, death, or external transfer. The study end date was standardized as August 25, 2020. The research protocol was approved by the institutional ethics committee (CAPPESQ: #4.107.580).

### Clinical data and outcome definitions

Data were obtained as part of the routine clinical care provided (routine-care-based cohort) and were extracted from patients’ electronic health records (EHR) onto standardized forms by trained extractors. Variables included sex, age, COVID-19 confirmation by reverse transcription polymerase chain reaction (RT-PCR) or serologic tests,[10, 11] underlying medical conditions reported by patients or relatives, procedures (mechanical ventilation, tracheostomy, dialysis or surgery), clinical outcome (discharge, death, external transfer or still hospitalized on August 25^th^) and patient itinerary (length of stay (LOS) in the emergency department (ED), hospital wards and ICUs for severe cases).[12]

### Cost analysis

The analysis was performed from the hospital’s perspective using micro-costing methodology.[13] Resources used by each patient were identified, quantified and valued to ascertain and describe individuals’ admission costs. Direct and fixed costs were included. Direct-cost subcategories of micro-costing for individuals’ admission costs included drugs, laboratory tests, radiologic exams, blood components and nutrition requirements. Direct costs of hospital supplies in the ED, wards and ICUs, including general supplies and personal protective equipment (PPE), were apportioned by patient-day in each hospital sector.

Other direct costs (human resources - medical staff and nonmedical staff) and the fixed costs (hospital services - laundry, food, administration, maintenance contracts, financing and general services) for each sector were apportioned by bed-day, resulting in daily medical staff, nonmedical staff and general daily cost estimates for each sector.

Indirect costs that were not related to the patient’s hospital admission (e.g., outpatient hospital visits, patient transportation, etc.) or the hospital’s functioning (e.g., productivity losses) were not included in this analysis.

The total admission cost for each patient was estimated by (i) the daily costs calculated according to the patient’s itinerary in the ED, wards and ICUs (PPE, supplies, medical staff, nonmedical staff, sector’s general daily cost) added to (ii) the direct costs measured by direct consumption (medication, laboratory tests, radiologic exams, nutrition and blood components). The total sector cost (ED, wards and ICUs) was calculated by summing all admission costs of each sector.

Costs were recorded in Brazilian Real (R$) and converted into US dollars (US$). From March 30^th^ to June 30^th^, US$1 was worth an average of R$5.55.

### Data analysis

Continuous variables are expressed as the mean ± standard deviation, and categorical variables are expressed as the number of cases and proportions. To assess the impact of different variables on hospital cost, the LOS of each patient subgroup was considered. An average cost (total cost/number of admissions) and a cost/day (total cost/total follow up in days) was estimated for each subgroup. Dividing each subgroup number of admissions by its total follow up, a rate (by 100 persons/days) was obtained. Dividing each subgroup total cost by this rate, we determine the cost/rate.

Missing data were treated as “no information” in the analyses, and they occurred only in comorbidities of 371 patients (11.4%) and SARS-CoV-2 RT-PCR results of 164 (5.0%) patients; no missing data occurred for the outcome variable. To fit multiple models, the cases with missing data for some variables were excluded.

The applied hypothesis tests considered an alpha error of 0.05. Continuous variables were compared among groups using Mann-Whitney U tests for independent samples. To obtain the adjusted impact of the interest variables on the total cost, a gamma generalized linear model (GLM) for y with a log link function was proposed. Statistical analyses were carried out using IBM SPSS Statistics v. 26.0 (SPSS Inc., Chicago, Illinois, USA) and R packages (R Core Team, Vienna, Austria).

## RESULTS

### Study population

Between March 30 and June 30, 2020, 3,254 admissions of patients with suspected or confirmed COVID-19, 54.5% of whom males, with an overall mean± standard deviation age of 58 (±18 years). COVID-19 was confirmed via RT-PCR or serologic tests in 2,512 (77.2%) patients, and the remainder were treated for presumed infections based on clinical and/or radiologic findings. Only 376 (11.6%) of the admissions were of patients with no comorbidities, and the others had 1 (23.2%), 2 or 3 (40%) or more than 3 (13.9%) comorbidities. The most frequent comorbidities were hypertension (48.1%), diabetes mellitus (30.5%), previous or current smoking (24.6%) and obesity (23%).

A total of 1,683 (51.7%) admissions comprised ICU stays. By August 25th, there were 2,016 (62%) discharges, 939 (28.9%) deaths, 278 (8.5%) transfers to other facilities and 21 (0.6%) patients remained hospitalized.

### Cost and follow-up

For the 3,254 patient admissions analyzed, there were 44,735 patient-days of hospitalization, resulting in a rate of 7.27 per 100 persons/day and a cost/rate of US$56,564.20. Figure 1a and b show the density of the follow-up time and the total cost (US$), including patient’s distribution. The total cost of the hospitalizations was US$41,122,173.39. The average cost per admission was US$12,637.42 (US$20,002.80 for the admissions that included ICU stays at any point, and US$4,839.57 for those that did not), and the overall daily cost was US$919.24.

**Figure 1.**
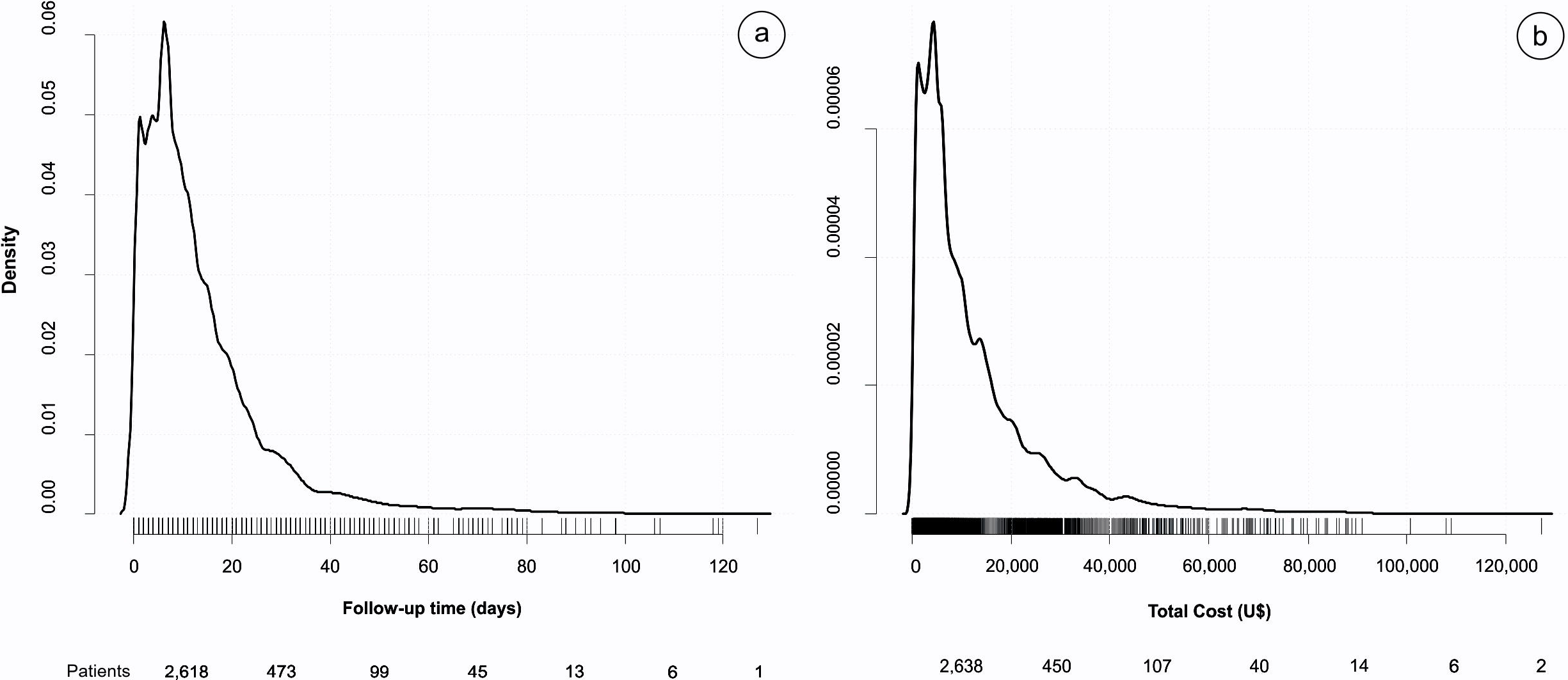
Density of the follow-up time in days and the total cost (US$).

The impact of age, sex, Covid-19 confirmation, comorbidities, procedures and outcome on hospital costs generally paralleled the total LOS experienced in patient-days (Table 1). Male patients were significantly more expensive (US$33,552.29 versus US$23,068.97 for females, p <0.001). The 55- to 65-year-old age group had the greatest impact on cost, with a mean US$17,791.12 cost/rate.

**Table 1.**
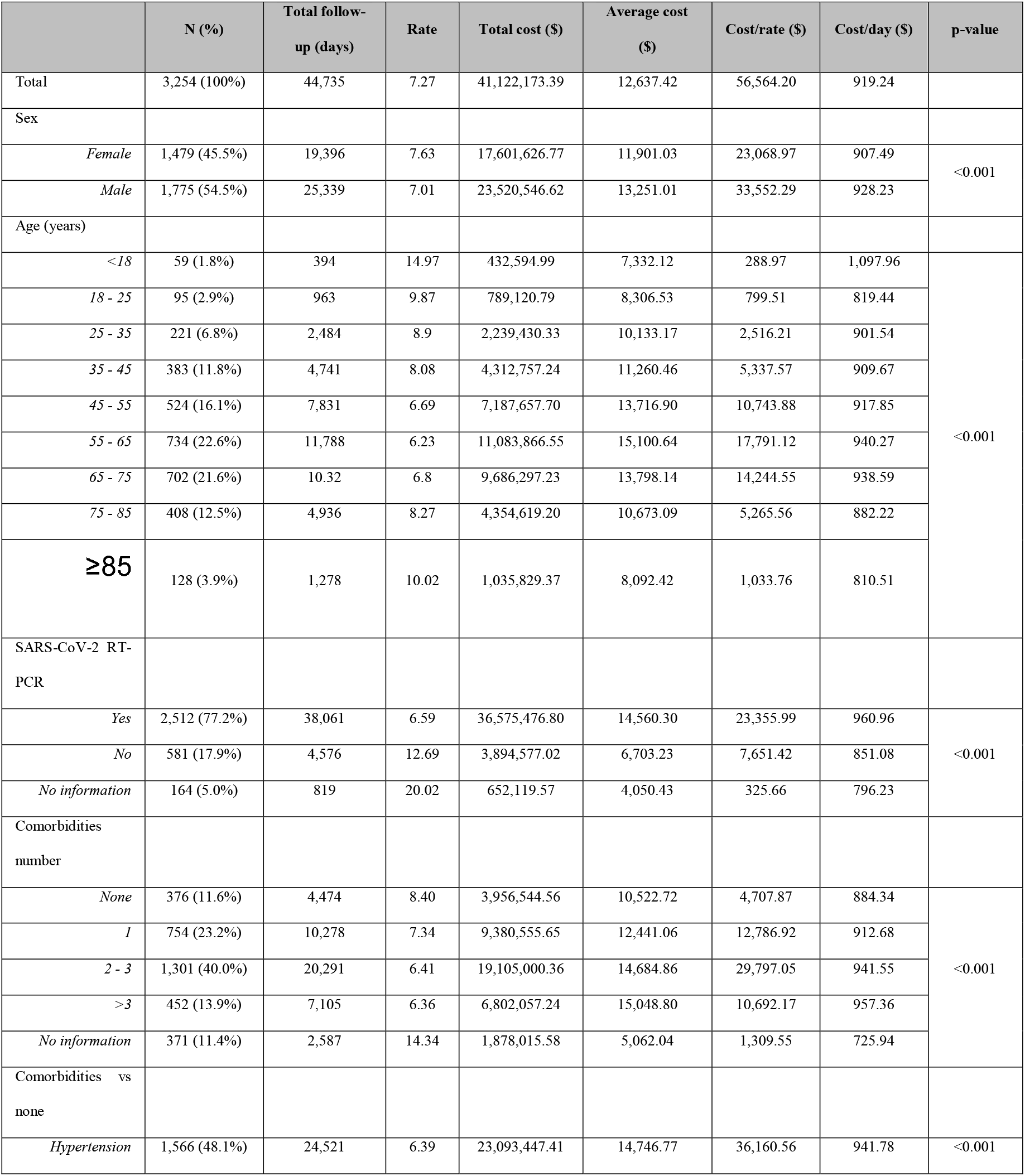
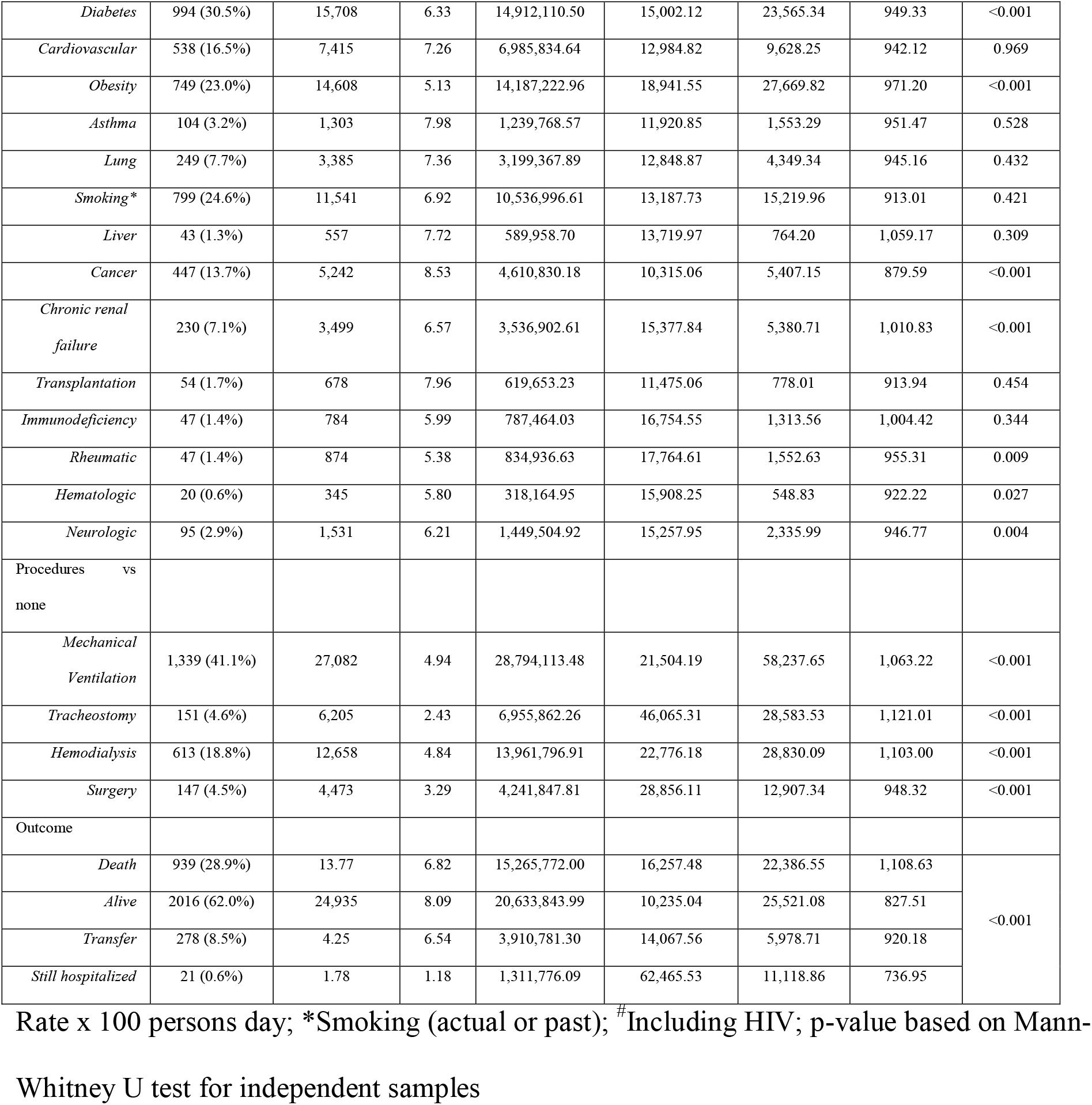
Descriptive Statistics, Including Absolute and Relative Frequencies, Total Follow-Up in Days, Rate Per 100 Persons/Day, Hospital Total Cost, Average Cost, Cost by Rate, and Cost by Day, According to Demographics, Clinical Conditions, and Status.

Admissions for patients with confirmed COVID-19 had higher average costs than those for patients without (US$14,560.30 versus US$6,703.23, respectively). Comorbidities significantly associated with higher average costs were hypertension (US$14,746.77), diabetes (US$15,002.12), obesity (US$18,941.55), cancer (US$10,315.06), chronic renal failure (US$15,377.84), rheumatic (US$17,764.61), hematologic (US$15,908.25) and neurologic diseases (US$15,257.95). The presence and number of comorbidities significantly increased the average daily cost according to a dose-response relationship; patients without comorbidities had a lower cost/rate (US$884.34/day) than patients with one comorbidity (US$ 912.68/day), 2 or 3 comorbidities (US$ 941.55/day) or more than 3 (US$ 957.36/day).

Requiring additional therapeutic procedures during hospitalization, i.e., mechanical ventilation (US$ 1,063.22/day), tracheostomy (US$ 1,121.01/day), hemodialysis (US$ 1,103.00/day) and surgery (US$ 948.32/day), were also significantly associated with higher average daily costs (overall US$919.24), compared to those that did not.

Age strata over 69 years (predicting 50% more costs) and 18-69 years (47% increase); laboratory-confirmed COVID-19 (61% higher costs); one (11%), 2 or 3 (19%) or more than 3 comorbidities (24%); and requiring mechanical ventilation (nearly 2 times more costs), dialysis (29%), surgery (79%), transfer (24%) or further hospitalization (97% higher costs) remained significantly associated with higher costs after adjustment by multiple regression analysis according to a gamma distribution. Costs of admissions with fatal outcomes were 24% lower than those with other outcomes in the adjusted analysis (Table 2). This model resulted in an overall US$1,562.8 rate difference (RD) (eFigure 1).

**Table 2.**
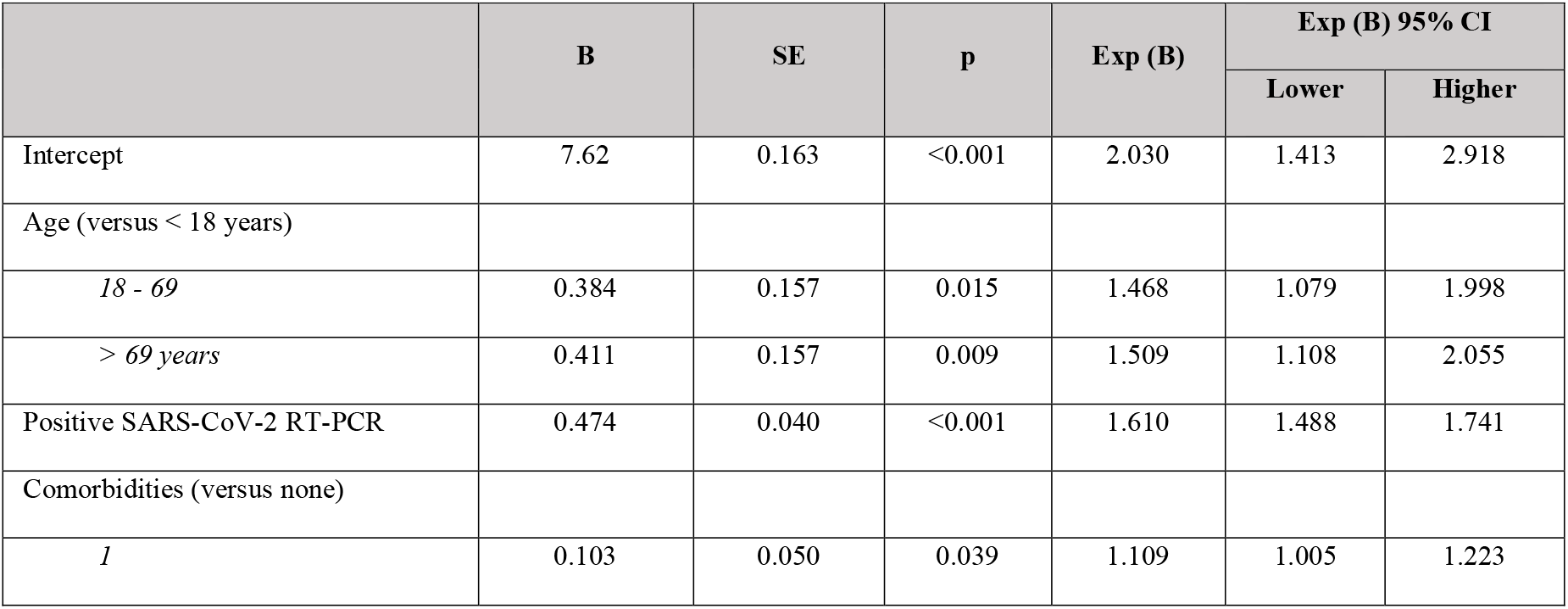
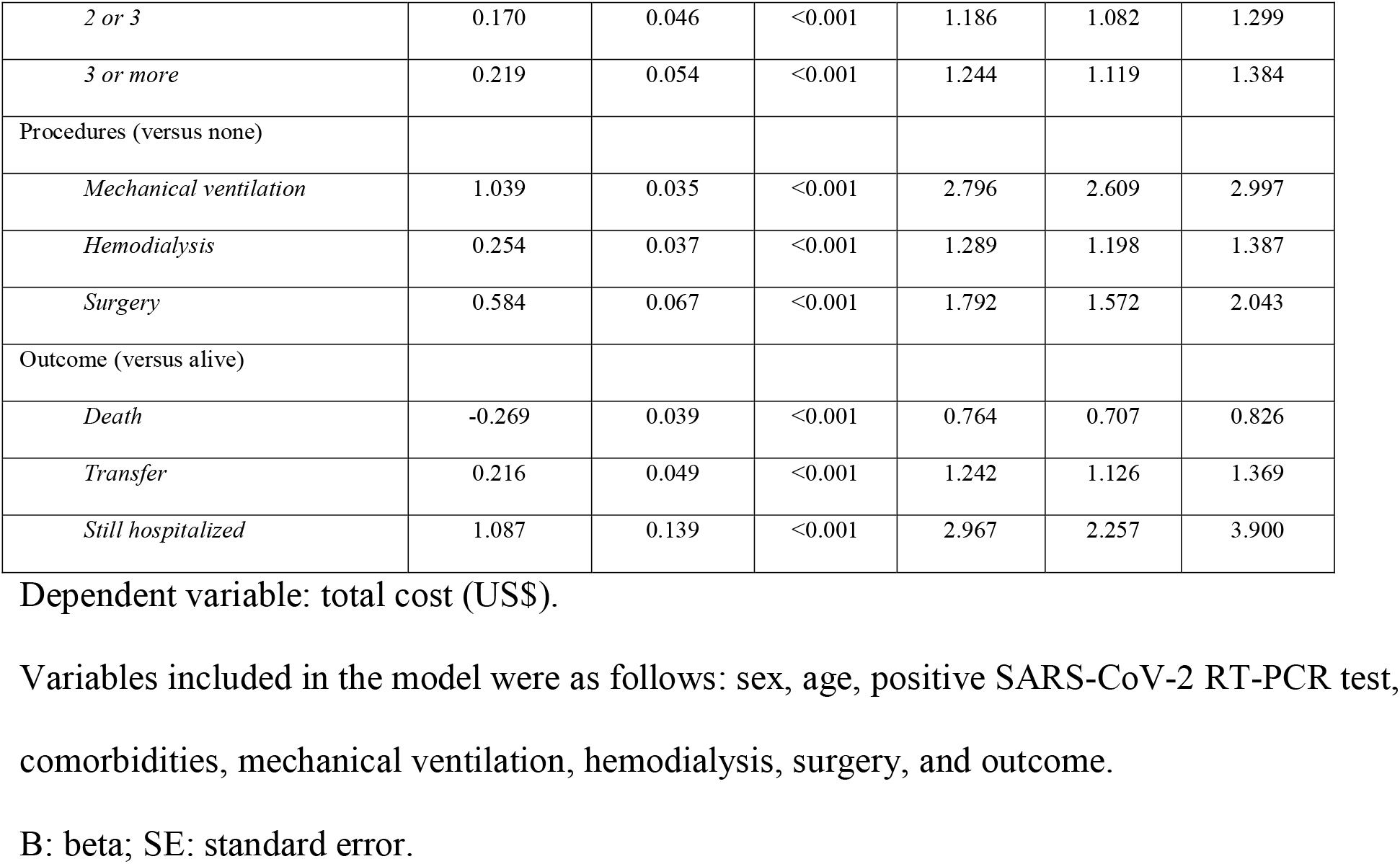
Multiple Regression Model with a Gamma Distribution to Assess the Adjusted Impact of Clinical Conditions on the Total Cost (US$).

|  | B | SE | p | Exp (B) | Exp (B) 95% CI |  |
| --- | --- | --- | --- | --- | --- | --- |
|  |  |  |  |  | Lower | Higher |
| Intercept | 7.62 | 0.163 | <0.001 | 2.030 | 1.413 | 2.918 |
| Age (versus < 18 years) |  |  |  |  |  |  |
| 18 - 69 | 0.384 | 0.157 | 0.015 | 1.468 | 1.079 | 1.998 |
| > 69 years | 0.411 | 0.157 | 0.009 | 1.509 | 1.108 | 2.055 |
| Positive SARS-CoV-2 RT-PCR | 0.474 | 0.040 | <0.001 | 1.610 | 1.488 | 1.741 |
| Comorbidities (versus none) |  |  |  |  |  |  |
| 1 | 0.103 | 0.050 | 0.039 | 1.109 | 1.005 | 1.223 |
| *2 or 3* | 0.170 | 0.046 | <0.001 | 1.186 | 1.082 | 1.299 |
| *3 or more* | 0.219 | 0.054 | <0.001 | 1.244 | 1.119 | 1.384 |
| Procedures (versus none) |  |  |  |  |  |  |
| *Mechanical ventilation* | 1.039 | 0.035 | <0.001 | 2.796 | 2.609 | 2.997 |
| *Hemodialysis* | 0.254 | 0.037 | <0.001 | 1.289 | 1.198 | 1.387 |
| *Surgery* | 0.584 | 0.067 | <0.001 | 1.792 | 1.572 | 2.043 |
| Outcome (versus alive) |  |  |  |  |  |  |
| *Death* | -0.269 | 0.039 | <0.001 | 0.764 | 0.707 | 0.826 |
| *Transfer* | 0.216 | 0.049 | <0.001 | 1.242 | 1.126 | 1.369 |
| *Still hospitalized* | 1.087 | 0.139 | <0.001 | 2.967 | 2.257 | 3.900 |

Dependent variable: total cost (US\$).
Variables included in the model were as follows: sex, age, positive SARS-CoV-2 RT-PCR test, comorbidities, mechanical ventilation, hemodialysis, surgery, and outcome.
B: beta; SE: standard error.

### Cost per sector

ICU stays had the highest costs (US$ 26,849,860.07; 64.7%), followed by ward (US$ 13,417,202.20; 32.3%) and ED (US$ 1,230,795; 92.3%) stays (Figure 2a). The overhead cost of nonmedical and medical staff was the component with the highest cost for all sectors (Figure 2b), representing 85% of the total admission costs in the ED, 82% in wards and 80% in the ICUs. In the ICUs, the other most important cost components were drugs (30.3%), supplies (23.3%) and laboratory tests (17.2%). In the wards, PPE, drugs and supplies had the highest costs (25.4%, 23.4% and 20.6%, respectively), and in the ED, they incurred by laboratory tests, radiologic exams and drugs (43.9%, 34% and 17.4%, respectively) (Figure 2c and d, Table 3).

**Table 3.**
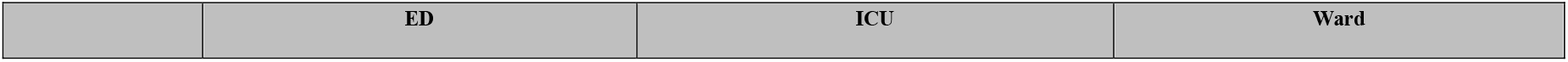
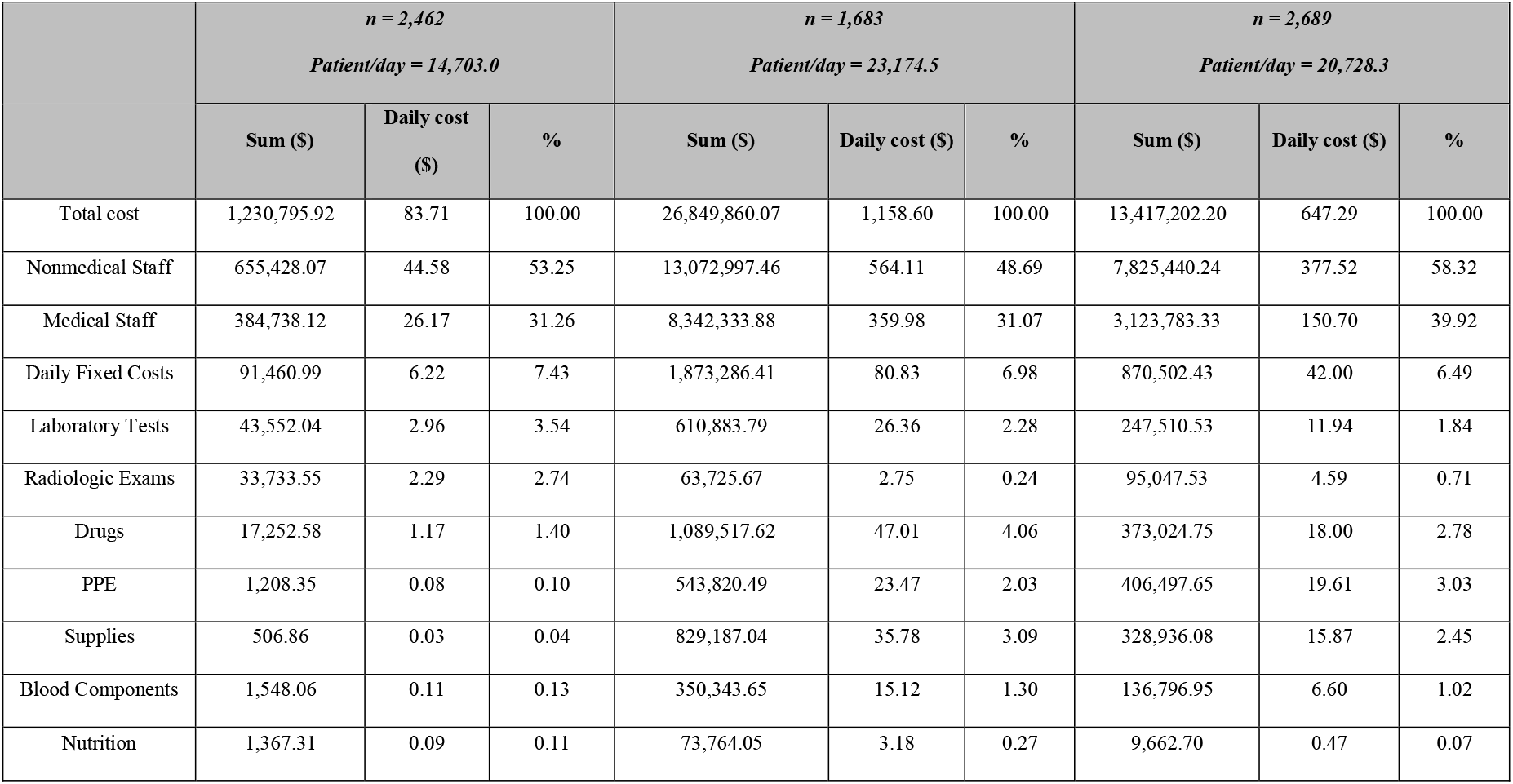
Cost Components and the Total Cost of Admission in Each Hospital Sector.

|  | *n = 2,462*<br>*Patient/day = 14,703.0* |  |  | *n = 1,683*<br>*Patient/day = 23,174.5* |  |  | *n = 2,689*<br>*Patient/day = 20,728.3* |  |  |
| --- | --- | --- | --- | --- | --- | --- | --- | --- | --- |
| | Sum (\$) | Daily cost (\$) | % | Sum (\$) | Daily cost (\$) | % | Sum (\$) | Daily cost (\$) | % |
| Total cost | 1,230,795.92 | 83.71 | 100.00 | 26,849,860.07 | 1,158.60 | 100.00 | 13,417,202.20 | 647.29 | 100.00 |
| Nonmedical Staff | 655,428.07 | 44.58 | 53.25 | 13,072,997.46 | 564.11 | 48.69 | 7,825,440.24 | 377.52 | 58.32 |
| Medical Staff | 384,738.12 | 26.17 | 31.26 | 8,342,333.88 | 359.98 | 31.07 | 3,123,783.33 | 150.70 | 39.92 |
| Daily Fixed Costs | 91,460.99 | 6.22 | 7.43 | 1,873,286.41 | 80.83 | 6.98 | 870,502.43 | 42.00 | 6.49 |
| Laboratory Tests | 43,552.04 | 2.96 | 3.54 | 610,883.79 | 26.36 | 2.28 | 247,510.53 | 11.94 | 1.84 |
| Radiologic Exams | 33,733.55 | 2.29 | 2.74 | 63,725.67 | 2.75 | 0.24 | 95,047.53 | 4.59 | 0.71 |
| Drugs | 17,252.58 | 1.17 | 1.40 | 1,089,517.62 | 47.01 | 4.06 | 373,024.75 | 18.00 | 2.78 |
| PPE | 1,208.35 | 0.08 | 0.10 | 543,820.49 | 23.47 | 2.03 | 406,497.65 | 19.61 | 3.03 |
| Supplies | 506.86 | 0.03 | 0.04 | 829,187.04 | 35.78 | 3.09 | 328,936.08 | 15.87 | 2.45 |
| Blood Components | 1,548.06 | 0.11 | 0.13 | 350,343.65 | 15.12 | 1.30 | 136,796.95 | 6.60 | 1.02 |
| Nutrition | 1,367.31 | 0.09 | 0.11 | 73,764.05 | 3.18 | 0.27 | 9,662.70 | 0.47 | 0.07 |

**Figure 2.**
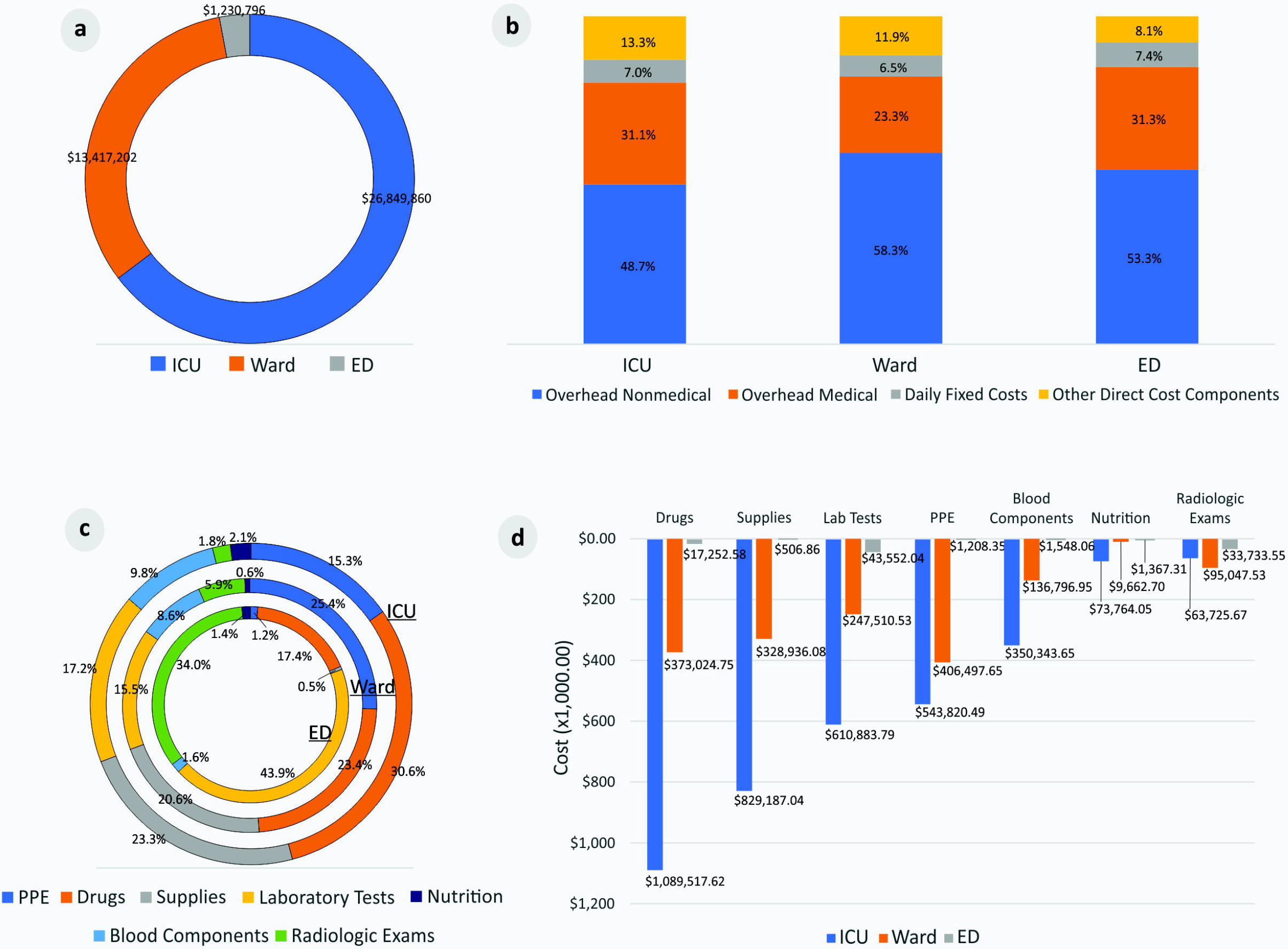
Cost components in ICUs, wards and the ED. (a) Total cost per hospital sector (US$), (b) % of cost components per sector, (c) % of other direct cost components in each sector, (d) Other cost components per sector (US$).

## DISCUSSION

Understanding the hospital costs of COVID-19 patients is essential to evaluating the economic impact of the pandemic on healthcare, providing important information for preparedness against and response planning for future risks, and improving knowledge regarding economic evaluation of global health emergencies.[1] Describing a large sample’s hospital costs according to patient clinical and demographic characteristics makes it possible to apply this analysis to different populations.

We report an average hospital admission cost of US$12,637.42, which is almost double China’s published cost of US$6,827 from 70 empirically observed cases[14] but is similar to the US$12,547 reported in a study from Saudi Arabia.[15] However, when interpreting average admission costs, the disease severity profile of the population must be considered. In our study, the average cost of admission was US$20,002.80 for the hospitalized severe cases admitted to ICU[12] (who accounted for 51.7%) and US$4,839.57 for the other cases. In a study of COVID-19 patients admitted to 2 hospitals in New York City, 22% of the patients were considered critically ill.[2] The population in our study resembles this group, since 41% required invasive mechanical ventilation, 19% required hemodialysis, and almost 29% resulted in death.[2, 3] This increased severity is due the existing risk-stratified healthcare network that was reinforced as part of the State’s COVID-19 pandemic response, with shelter hospitals for mild cases’ hospitalizations and referral units, like HCFMUSP, were designated for patients with severe conditions. So, regulators transferred critically ill COVID-19 patients and those with complex underlying conditions requiring specialized care to HCFMUSP, where the ICU capacity was increased.[5, 6] As expected, in our study, the highest cost/day was incurred during the ICU stay.

The correlation between pre-existing health conditions, age and disease severity was established previously.[16, 17] Here, we describe how these variables are also related to higher costs. An increasing trend was observed with the number of comorbidities, where compared to no comorbidities (US$10,522.72), 2 or 3 comorbidities (n= 1,301) elevated the average admission cost by 16%, and >3 comorbidities (n= 452) increased it by 19%. These findings are also similar to Shandong’s report, with a 40% increase in hospital admission cost for patients with any pre-existing disease.[14] Laboratory confirmation of SARS-CoV-2 infection was also independently associated with higher admission costs and may be related to disease severity, with a higher viral load being present in more severe cases.[18, 19] Death was the admission outcome related to lower costs, probably due to limiting the LOS by occurring early in the disease course (eFigure 2).

Human resources accounted for the greatest proportion of costs in all hospital sectors. In the ED, this component substantially elevated the cost/day, which can be partly explained by the ED needing to have medical and surgical specialties available 24/7 to assist with the specific urgent needs of inpatients from all sectors. The COVID-19 pandemic response increased the need for healthcare professionals since they were essential to capacity expansion. Competitive salaries had to be paid given the limited number of professionals, and this required a 46% increase in shift wages, which also increased overhead costs.[20]

Analyzing the costs of COVID-19 patient admissions and comparing them to costs incurred during other epidemics puts the costs of COVID-19 hospital admissions into a historical context. The average hospital admission cost of US$12,637.42 is similar to the model means of US$12,264, which was predicted using data for severe pneumonia hospitalizations due to H1N1 in the USA,[21] and US$12,947, which was based on MERS-CoV data from Saudi Arabia.[15] The reported hospital costs of seasonal influenza infections are higher; a study described them to be US$34,743 for patients aged between 20 and 64 years.[22] Data about other epidemics showed that the cost of treatment of patients with pertussis ranged from $412–$555[23] and estimated the cost for treating 11 patients with Ebola virus disease during the 2014 outbreak in West Africa to be US$1.2 million in a treatment center in the USA.[24, 25]

Economic evaluations are usually impacted by local conditions related to resource availability and the local market. However, in the COVID-19 pandemic, some aspects were due to global concerns, such as the simultaneous worldwide need for PPE and mechanical ventilators.[26] Our study reinforces the impact of PPE on costs, as they totaled US$951,526.50.

In 2009, The National Institute for Health and Care Excellence (NICE) set the nominal cost-per-QALY threshold at £50,000 for end-of-life care.[27] Therefore, considering the effectiveness in saving lives of the support given by HCFMUSP to COVID-19 patients, compared to no lives being saved, the average cost of US$12,637.42 described in the present study seems reasonable. The costs of COVID-19 sequelae and their social and economic impact should also be analyzed in future studies.

This study has some limitations: it took place in a single quaternary reference center and was performed from the hospital perspective, excluding indirect costs and additional investment in fixed capacity expansion and equipment acquisition. However, the study findings were analyzed with established, standardized, reproducible methods with the aim of supporting referral hospitals’ emergency preparedness. Therefore, the prospective collection of detailed clinical data, sector itineraries, outcomes, resources used and costs for a large sample of patient hospitalizations are strengths of this study.

## CONCLUSION

In this single quaternary reference center, human resources accounted for the largest cost component, and ICU for the costliest sector. Total costs and average and daily estimated costs increased by 50% for older age strata, by 10-24% according to the number of comorbidities and by 24%-200% when additional therapeutic procedures were required; these costs decreased by 24% when the outcome was death.

Understanding the hospital costs related to COVID-19 admission and the economic impact of the disease across different population subgroups can support the development of a comprehensive approach to hospital preparedness, decision-making and planning for future risk management, and is the first step in evaluating the cost-effectiveness of treatments and vaccination programs.

## Data Availability

Deidentified participant data and a data dictionary defining each field in the set will be made available on request. They will be sent by the corresponding author after approval of a proposal with a signed data access agreement. Data will be available on publication.

## ACKNOWLEDGMENTS

EPICCoV Study Group, COVID Registry Group, Alessandra Pereira, Francis Mironescu Tomazini, Claudia Maria Montebello de Oliveira, Givaldo Oliveira de Souza, Ligia Maria Dal Secco.

## AUTHOR CONTRIBUTIONS

AMM, AJFC, EvT, LACD, and LH conceived and designed the study; AMM, AJFC, HP, ErT, VC, FARJ, RASL, EvT, and LH participated in data acquisition and analysis; AMM, AJFC, HP, EvT, LACD, and LH participated in the interpretation of results; AMM, AJFC, HP, EvT, and LH drafted the article. All authors read and approved the final manuscript.

