## supplementary file for "Unraveling COVID-19-related hospital costs: The impact of clinical and demographic conditions"

**Miethke-Morais et al. 2020.**

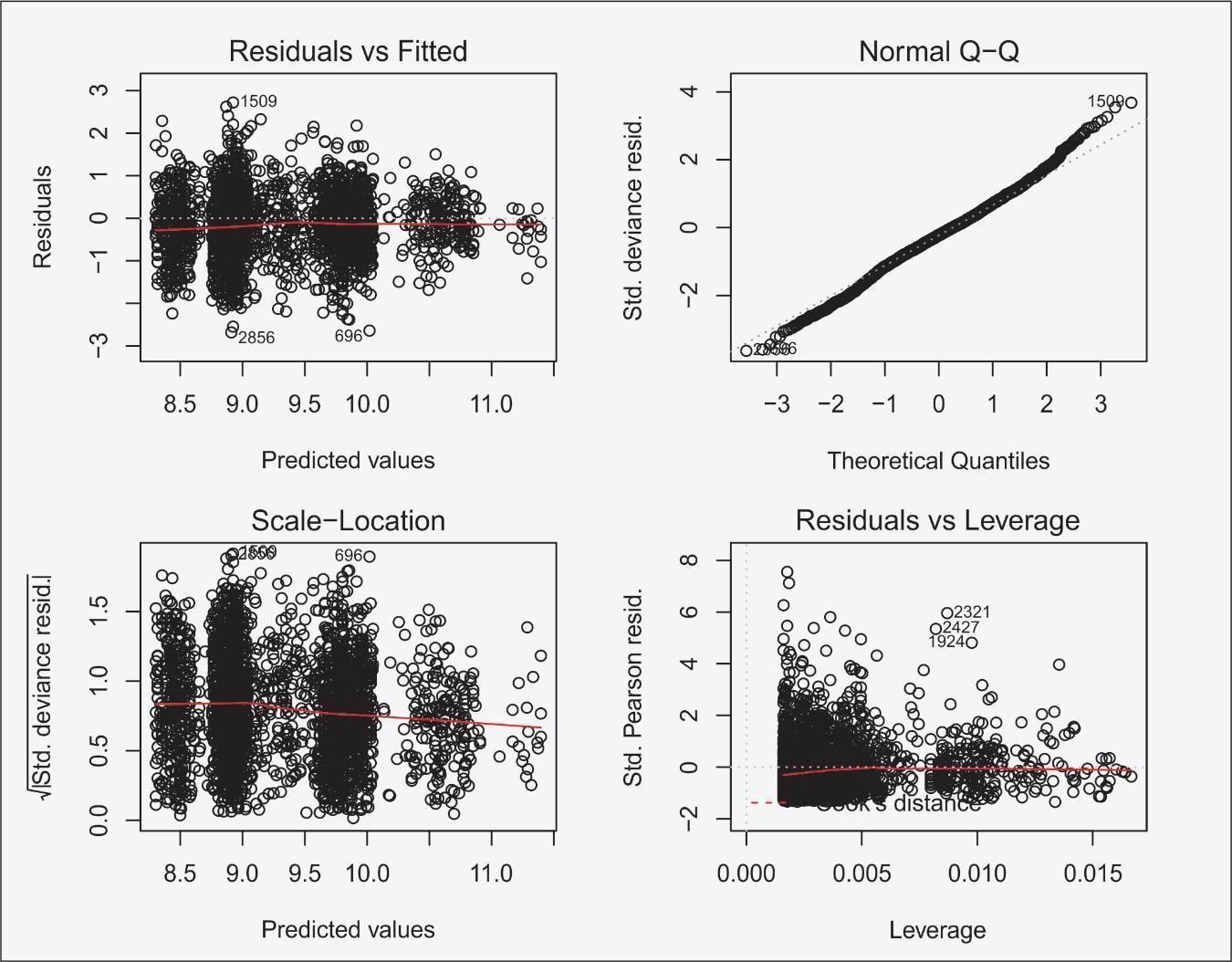

**Supplementary Figure 1.** Basic diagnostic plot of the multiple gamma generalized linear models (GLMs) for y with a log link function

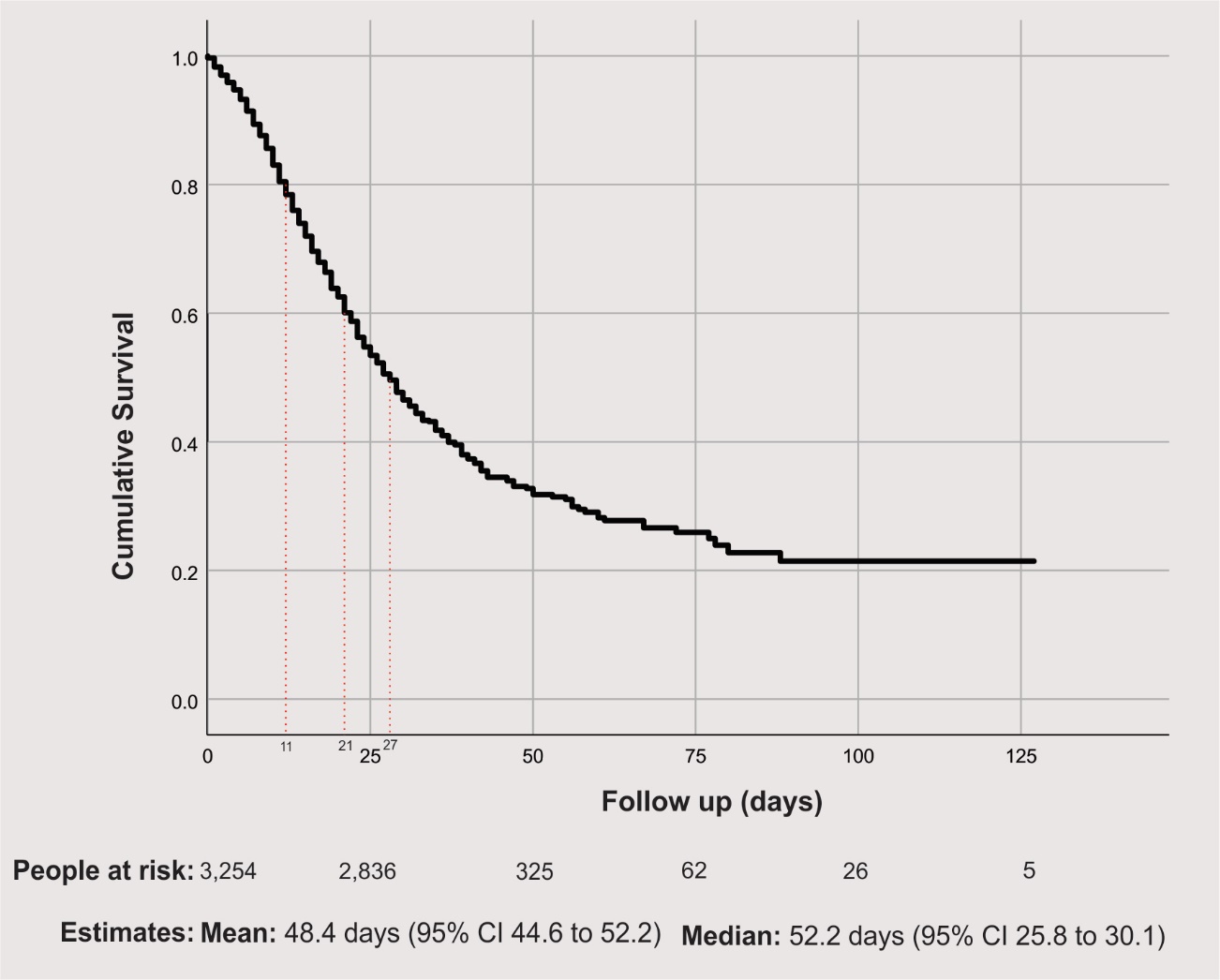

**Supplementary Figure 2.** General survival, where dotted red lines indicate a 20%, 40% and 50% risk of death
